# Nutrition, Cognition and Learning: The Role of White Matter Development and Connectivity

**DOI:** 10.64898/2026.07.01.26356557

**Authors:** Diandra Brkić, Jonas Hauser, Charles-Edouard Rouault, Iaroslava Semenova, Fabio Mainardi, Sean Deoni

## Abstract

While substantial research has examined the role of infant nutrition in early brain and cognitive development, the links between later childhood nutrition, brain development, and cognitive and academic skills remain less explored. In this work, we investigated for the first time the direct and indirect associations between nutrition intake, white matter microstructure and structural connectivity, and cognitive and academic outcomes. Using longitudinal data from typically developing children aged 2 to 14 years, we combined neuroimaging, dietary, and cognitive measures to test direct associations between nutrition and brain structure and connectivity, as well as cognitive and learning outcomes, and to assess whether brain development mediated these relationships. We found that specific nutrients, including DHA, sphingomyelin, iron, niacin, choline, and palmitoleic acid (omega-7), were associated with improved brain structure and connectivity, as well as better cognitive and learning outcomes. We also found that brain development partly mediated the association between childhood nutrition and learning outcomes. To our knowledge, this is the first study to show such a pathway in school-age children. These results add to the growing literature demonstrating the ongoing importance of nutrition beyond infancy in supporting childhood brain and cognitive development.

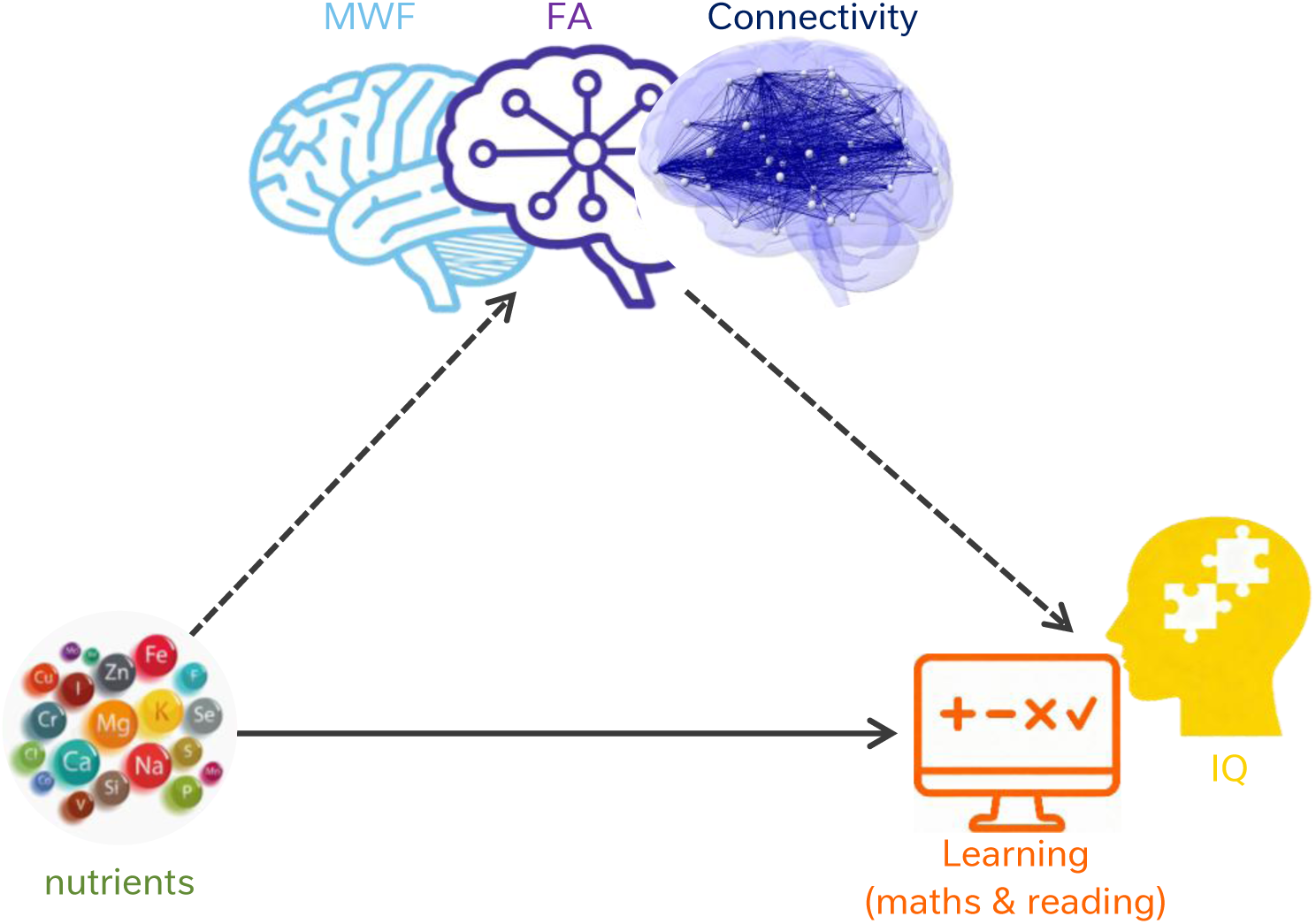

## INTRODUCTION

Childhood, roughly between ages 5 and 13 years, is a period of rapid neurodevelopment marked by dynamic brain changes that support cognitive development (1–3). Visuomotor, language, and emotional skills established in the first 1,000 days provide the foundation for more complex cognitive abilities that emerge during childhood. These include executive functions, socioemotional processing, and other academic-related skills such as reading and maths (1,4). Underpinning this cognitive maturation is the refinement of the brain’s eloquent functional systems and networks (5). In particular, global and regional morphological and functional changes accompany key developmental milestones and transitions in cognition and behaviour that shape future and long-term outcomes (6–8).

Although neurodevelopmental processes like myelination, synaptogenesis, and subsequent pruning, have markedly slowed from their peak rates in the late foetal period and infancy, these processes continue throughout childhood into young adulthood. Resultant differences in brain morphometry and white matter microstructure and architecture have been consistently associated with developing cognitive abilities, such as processing speed, executive functioning (EF), motor learning, attention, working memory, and learning in both typical and atypical developing groups. Thus, although much of the past research focused on neurodevelopment has prioritised the early infant period, less-than-optimal brain development throughout childhood can also have lasting negative consequences on education, career opportunities, and mental well-being (13).

Multiple biological and environmental factors contribute to successful brain development and educational outcomes. Amongst the most consistently identified predictors are socioeconomic factors (including parental education, income), the home environment (including aspects of inter-partner violence, stress, and supportive caregiving), maternal and child health factors (including preterm birth, birth weight, gestational age, and exposure to neurotoxicants), and nutrition (14). Due to the diverse and nonlinear spatiotemporal pattern of neurodevelopment, the timing and magnitude of social, biological, and environmental factors may lead to differing outcomes. With respect to nutrition, evidence from both cross-sectional and longitudinal studies suggest general malnutrition as well as specific nutrient deficiencies throughout childhood can have lasting negative impacts on neurodevelopment, educational outcomes, and later success in life (15,16). These findings have promoted extensive public health initiatives that support infant and childhood nutrition and education.

*Nutritional cognitive neuroscience* seeks to understand how dietary patterns and specific nutrients shape brain structure, function, and cognition across the lifespan (17). Like developmental neuroscience in general, the vast majority of past nutritional cognitive neuroscience research has focused on early nutrition, specifically infant nutrition and breastfeeding (18). In contrast, studies of later child and adolescent development have predominately examined relationships between food insecurity or suboptimal dietary habits and academic performance (19). For instance, there is a relationship between frequency of consumption of certain foods (e.g. diary, meat, eggs) and academic achievement (20), micronutrient deficiencies (i.e. iron) and poor school performance (21), or unhealthy nutritional habits (e.g., skipping breakfast) and school performance (22). Few studies have directly explored the interplay between nutritional intake, brain development and/or function, and cognitive performance in typically developing children and young adolescents, and even fewer have combined neuroimaging, detailed dietary assessment, and direct cognitive testing. Recent systematic evidence suggests that diet quality (e.g., Mediterranean) and specific nutritional components, such as lipids, are positively associated with neural and cognitive development, although methodological heterogeneity and the limited number of studies available constrain definitive conclusions (18,19).

The present study aims to address this gap by leveraging longitudinal data from the RESONANCE study of child health and development. This study, initiated at Brown University in Rhode Island, USA, in 2009, has longitudinally followed ∼1800 children from foetal and early infancy through to middle adolescence with repeated assessments of brain structure and function (MRI neuroimaging), cognitive development (using standardized scales and tools), genetics, environmental factors, social determinants of health, and nutrition. Building upon prior evidence from cohorts of younger children demonstrating associations between nutritional components and brain areas supporting specific cognitive outcomes (23), we first were interested to check whether these associations would be relevant in childhood, and second to study whether white matter characteristics (myelination and fractional anisotropy) mediate the relationship between nutritional components and cognitive and learning outcomes, specifically IQ, math and reading abilities. We hypothesised that nutritional components previously highlighted in infancy (e.g., docosahexaenoic acid (DHA), iron, choline, sphingomyelin) would continue to be of importance at school age (24).

## METHODS

### Study Design & Participants

This study included a subset of 282 (115 females) healthy, full-term, and neurotypically-developing children (mean age of 7.4 years) who were drawn from the Brown University [RESONANCE study](https://atlaslongitudinaldatasets.ac.uk/datasets/echo-resonance) (Table 1). We included only the participants who had completed the nutritional assessment. More specifically, each child in full RESONANCE cohort was recruited prior to age 5 and followed at approximate 6-month intervals between birth and 2 years of age, and 12-month intervals from 2 through adolescence. To focus on neurotypical development children were excluded if they were or had: born pre-term (less than 37 weeks); small for gestational age (<1500g); non-singleton pregnancy; complicated delivery requiring a stay in the NICU or 5 minute APGAR scores less than 8; *in utero* exposure to cigarettes, alcohol, or illicit substances; family history of learning or psychiatric disorders (including major depressive disorder in the mother requiring medication during pregnancy); or a history of neurological trauma or disorder (e.g., epilepsy). For this analysis, children between 2 and 14 years were selected who had a combination of brain imaging (either relaxometry or diffusion imaging), nutrition-recall (ASA24) data, and had completed neurocognitive testing as described below. Written informed consent was obtained from each child’s parents or legal guardian, and the study was performed with approval from the institutional internal review board.

**Table 1.** **Demographic description** of the data sample used in this study. Note that some participants had both WISC and WPPSI measured at different time points. For the analysis we have included the composite FISQ score.

| Demographic | N=282 |
| --- | --- |
| **Age** |  |
| Mean (SD) | 7.41 (2.44) |
| Median [Min, Max] | 7.27 [2.77, 14.3] |
| **Sex** |  |
| Female | 115 (40.8%) |
| Male | 167 (59.2%) |
| **Ethnicity** |  |
| Hispanic/Latino | 41 (14.5%) |
| Non-Latino/Hispanic | 233 (82.6%) |
| Unknown | 2 (0.7%) |
| Missing | 6 (2.1%) |
| **WISC IQ** |  |
| Mean (SD) | 111 (15.5) |
| Median [Min, Max] | 113 [62.0, 141] |
| Missing | 52 (18.4%) |
| **WWPSI IQ** |  |
| Mean (SD) | 109 (13.1) |
| Median [Min, Max] | 111 [69.0, 136] |
| Missing | 186 (66.0%) |
| **Reading (AAB)** |  |
| Mean (SD) | 115 (23.4) |
| Median [Min, Max] | 118 [50.0, 150] |
| Missing | 52 (18.4%) |
| **Maths (AAB)** |  |
| Mean (SD) | 108 (19.0) |
| Median [Min, Max] | 108 [50.0, 150] |
| Missing | 51 (18.1%) |

### Data Collection Methods

#### MRI Imaging

Neuroimaging was performed on a Siemens Trio 3T with a 12-channel head RF array. Children under 4 years of age were typically imaged during natural non-sedated sleep (25). To minimize intra-scan motion, children were swaddled with an infant or pediatric MedVac vacuum immobilization bag (CFI Medical Solutions, USA) and foam cushions were packed around their head. Scanner noise was reduced by limiting the peak gradient amplitudes and slew-rates to 25 mT/m/s. A noise-insulating insert (Quiet Barrier HD Composite, UltraBarrier, USA) was also fitted to the inside of the scanner bore. MiniMuff pediatric ear protectors and electrodynamic headphones (MR Confon, Germany) were used for all children. Older children and those able to tolerate awake scanning were imaged whilst watching a movie or TV show and were also compressed within a pediatric MedVac vacuum immobilization bag, and inflatable (PearlTec) and memory foam cushions used to minimize body and head motion. A pediatric pulse-oximetry system and infrared camera were used to continuously monitor the infants and children during scanning, and parents were allowed to sit with their child in the scanner suite provided they had no MRI counter indications.

To provide general information on brain structure and organization, including cortical, and sub-cortical morphometry, a high spatial resolution (1×1×1) mm^3^ T_1_-weighted magnetization-prepared gradient recalled echo (MP-RAGE) image was acquired.

To assess white matter development and myelination, mcDESPOT multicomponent relaxometry and diffusion tensor imaging (DTI) were performed. mcDESPOT (26,27) provides voxel-wise estimates of the myelin water fraction (MWF), a quantitative and surrogate measure of myelin content (28,29). Multicomponent relaxometry exploits the inherent sensitivity of T_1_ and T_2_ relaxivity to biochemical structure and composition to resolve sub-voxel water species. In human brain, two such species are consistently observed, a slow-relaxing water pool attributed to intra/inter-cellular water, and a fast-relaxing water pool attributed to water trapped within the lipid bilayers of the myelin sheath (28). The ratio of the myelin to the non-myelin water pool, termed the myelin water fraction or MWF, provides and non-invasive and quantitative estimate of myelin content.

Multiple b-shell diffusion imaging data was acquired using a twice-refocused echo planar imaging (EPI) protocol amenable to both conventional diffusion tensor and non-tensor analysis. 32 non-collinear diffusion encoded images were acquired at 3 incremented diffusion weightings (b values) of 700, 1500, and 2200 s/mm^2^ along with 5 reference non-diffusion-weighted b=0 images.

#### Neurocognitive assessments and sociodemographic data collection

Within seven days of successful scanning, children return to the lab for an in-person cognitive assessment that included both observational measures Wechsler Intelligence Scale for Children – Fifth Edition, WISC-V (30), the Wechsler Preschool and Primary Scale of Intelligence (WPPSI-IV) for preschool children (31), and the Academic Achievement Battery, AAB, for children older than 4 years (32), parent report (the Behavior Rating Inventory of Executive Function, BRIEF) and self-administered assessments (The NIH Toolbox) (33,34). In addition, child behavior screening tools, including the Modified Checklist for Autism (M-CHAT) and the Child Behavior Checklist (CBCL) were used to identify children with potentially clinically worrisome behaviors and remove them from the normative child cohort (35,36). Here we focussed our analysis on the cognitive (WPPSI and WISC-V) and learning (AAB) outcomes.

Extensive infant, parent, and family health and sociodemographic information was also collected on each child and family. This included neurological and psychiatric history, maternal and paternal education levels; maternal prenatal and postnatal health, substance use, breastfeeding practices, gestation duration, and birth weight.

From the set of administered forms, the following composite measures were used for the analysis presented herein: the word/letter recognition (WLR) and mathematic calculation (MC) domains from the AAB; full-scale intelligence quotient (FSIQ) from the WISC-V and the WPPSI-IV.

At the same visit as the neurocognitive assessments, parents were asked to complete a series of questionnaires that provided an overview of family demographic and household characteristics (e.g., race, ethnicity, marital status, family size and composition, household income, parental education levels, and employment status and profession), as well as maternal and child health history, including pregnancy complications, gestation, birth length and weight, APGAR scores, and breastfeeding and infant feeding history.

#### Nutritional intake assessment

The Automated Self-Administered 24-Hour Recall (ASA24) was used to assess nutritional intake. The ASA24 is a web-based questionnaire that collects detailed information about foods and beverages consumed over a 24-h period. Parents were asked to recall all the foods and beverages consumed by their child each day for at least three separate days (including at least one weekend and two weekdays).

Nutrient intakes were automatically derived from the ASA24 reports using Food and Nutrient Database for Dietary Studies ([FNDDS](https://www.ars.usda.gov/nea/bhnrc/fsrg/)), and food group intakes were derived using the USDA Food Patterns Equivalents Database ([FPED](https://www.ars.usda.gov/nea/bhnrc/fsrg/)).Output variables included total energy intake (kcal/day), macronutrients (e.g., carbohydrate, protein, total fat), micronutrients (e.g., vitamins and minerals) expressed in absolute amounts and/or density-adjusted units as appropriate. In addition, values for amino acids, were obtained from standard USDA food composition databases ([FoodData Central](https://fdc.nal.usda.gov/)). Values for choline-containing phospholipids, including phosphatidylcholine and sphingomyelin, were obtained from the USDA Database for the Choline Content of Common Foods (37).

### MR Image Analysis

#### Myelin Water & Relaxometry Processing

Following image acquisition, the mcDESPOT multicomponent relaxometry data were visually assessed for motion artifacts (e.g., blurring and ghosting) by the same researcher (SCLD) and MWF myelin content measures estimated throughout the brain on a voxel-by-voxel basis using a 3-pool tissue model. The quantitative MWF images were non-linearly aligned to MNI space using a multi-step, multi-scale approach as previously described (38). Brain regions with similar myelination patterns have previously been identified by merging 1290 longitudinal MWF images from 587 (259 females) healthy and neurotypically-developing children from 45 days to 13.4 years of age. Probabilistic independent component analyses (PICA) was used to delineate non-orthogonal but statistically independent spatial regions with shared temporal myelination trajectories in an unsupervised and data-driven manner (39). This analysis provided 176 independent components (ICs) derived from the estimate of Bayesian evidence, which were used as myelination region-of-interest (ROI) masks.

#### Diffusion Tensor Analysis and Structural Connectivity Processing

From the collected diffusion weighted images, a standard processing pipeline was used that included eddy current correction and b-matrix rotation, followed by voxel-wise tensor fitting and fibre orientation distribution estimation using FSL’s DTIFit and BEDPOSTX, respectively. Quantified fractional anisotropic (FA) maps were non-linearly aligned to MNI space a similar approach as applied to the MWF images, with the b=0 T_2_-weighted image first aligned to the MNI template, and the registration matrix then applied to the FA maps.

To quantify structural connectivity, each child’s T_1_-weighted MP-RAGE image was run through Freesurfer (version 7.4) to segment the cortical ribbon and delineate the cortex into 34 regions per hemisphere, as well as subcortical deep grey matter structures. At each stage throughout the processing pipeline, images were visually inspected and, if needed, manually edited and corrected. This included inspecting data for poor skull-stripping, which required re-running this processing step, the additional use of gcut and, in extreme cases, manual removal of the remaining dura and eye signal. The segmented region masks were then non-linearly aligned to the child’s non-diffusion-weighted b=0 image as used as seed and target regions for probabilistic tractography (probtrackX). Tractography was initiated from each cortical seed mask, with all remaining cortical masks serving as targets. From each seed voxel, 5000 streamline samples were generated by repeatedly sampling from the local posterior fibre orientation distributions estimated by BEDPOSTX. Streamlines were propagated using a fixed step length (0.5mm) and curvature threshold (0.2). Tracking was constrained to a whole-brain white matter mask with an FA threshold of 0.1. Streamlines were terminated upon reaching target masks, exiting the brain mask, or violating curvature constraints.

Lastly, connectivity between each pair of seed and target masks was quantified as the number of successfully propagated samples from the seed region that reached each target region. To account for differences in ROI size and total number of generated samples, connectivity values were normalized by the total number of samples initiated from the seed region, yielding a measure of connection probability. This procedure resulted in a weighted structural connectivity matrix for each child, representing pairwise probabilistic connectivity between cortical and sub-cortical regions.

#### Brain, Cognition, and Nutrition Modelling

The following sets of analyses were performed to examine the direct associations between nutrient intake (ASA24) and brain white matter development (assessed with MWF, FA, and structural connectivity), and white matter development and cognitive function (AAB, MC and LWR, and FSIQ). Finally, a mediation analysis was performed to quantify the direct effect of nutritional status on cognitive performance and the indirect effect mediated by white matter development, with significance of indirect effects assessed using bootstrapped confidence intervals.

#### Associations Between Nutrient Intake and White Matter Development

We investigated the association between nutrition and myelination patterns in the 176 myelination ROIs by fitting a series of linear mixed-effects models that included MWF or FA as the outcome measure and nutrient intake (NI), total calorie intake (TotCal), log(child age), biological sex, and maternal education (MatEd) as predictor variables, i.e.,

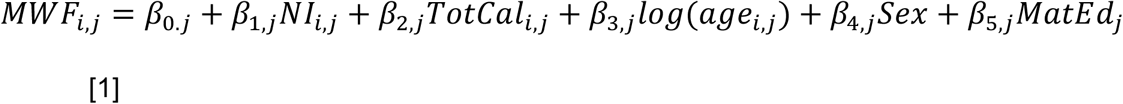

Or

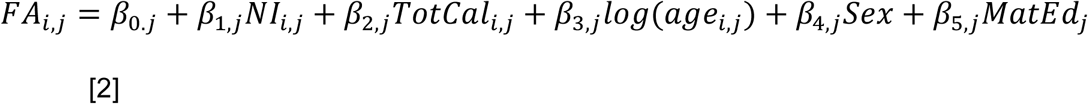

where, *MWF_i,j_* or *FA_i,j_* is the mean MWF or FA estimate within an ROI of child *j* at time-point *i*, *age_i,j_* is the corresponding child age, *β_0,j_* is the intercept and *β_1,j_ to β_5,j_* are the regression coefficients that combine a sample fixed effect and a subject-specific random effect.

From this analysis, regions with a p value for β_1_ < 0.001 were identified and merged into a single mask. Mean myelination within this mask was then calculated for each child and timepoint, and the model rerun. Nutrients with β_1_ < 0.05 (FDR) were then determined.

To reduce the number of nutrients tested, we restricted our analysis to nutrients with known or potential associations to brain development, including: saturated fatty acids (e.g, hexadecanoic acid), mononsaturated fatty acids (e.g., octadecenoic acid), and polyunsaturated fatty acids (including docosahexaenoic and arachidonic acids), as well as cholesterol, sphingomyelin, choline, folate, vitamins B6, B9, B12, C, and D, iron, iodine, zinc, magnesium, niacin, and folic acid).

#### Associations Between Nutrition Intake and Cognitive Development

To assess relationships between nutrition intake and the FSIQ and AAB LWR and MC cognitive scores we used the mixed-effects model,

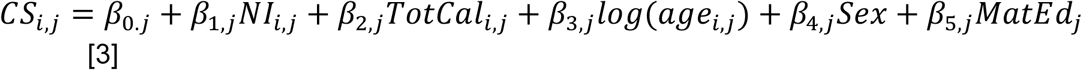

#### Mediation Analysis linking Nutrition, Myelination, and Cognitive Development

Finally, mediation analysis was performed using the results from the prior sets of association analyses. Specifically, we tested whether brain myelination mediates the relationship between nutrition and cognitive performance. Using the CANlab mediationToolbox (40), for each nutrient-cognitive score paring we specified a single mediator model with nutrient intake as the predictor, X, cognitive score as outcome, Y, and brain region myelin as the mediator, M. We included child biological sex, maternal education, log(child age), and total calorie intake as covariates. The model estimated the canonical mediation paths: a (X→M), b (M→Y controlling X), c (total X→Y), and c′ (direct X→Y controlling M), and the indirect effect as the product a·b. Statistical inference for the direct and indirect effects used the toolbox’s bootstrap-based testing and significance was evaluated using two-sided bootstrap p-values for the direct (c′) and indirect (a·b) effects. From this analysis, brain regions with an indirect path p value < 0.001 were identified and merged into a single mask. Mean myelination (MWF or FA) within this mask was then calculated for each child and timepoint, and the model rerun.

#### Mediation analysis linking Brain Connectivity, Nutrition, and Cognitive Development

Building on the analysis of white matter microstructure, we next assessed the direct and indirect associations between measures of nutrient intake, brain connectivity, and cognitive performance using the mediation framework described in 2.4.3. The indirect path p value matrix (i.e., the p value associated with each pairwise connectivity measure) was constructed and connectivity hubs were identified using degree centrality and strength.

## RESULTS

### Associations Between Nutrient Intake and White Matter Development

Results from our analysis investigating relationships between nutrient intake and white matter development are summarized in **Figures 1** and **2** for MWF and FA, respectively. Nutrients showing positive associations, i.e. greater intake was associated with increased MWF, included choline, docosapentaenoic acid (DPA), hexadecanoic acid (palmitic acid), magnesium, niacin, and sphingomyelin. Most of these nutrients have previously been associated with neurodevelopment generally (i.e. myelination), and white matter development more specifically. Prior work shows how long-chain fatty acids, choline, phospholipids, and sphingomyelin are implicated in myelin formation and maintenance. Sphingomyelin is a major myelin-associated phospholipid, considered a structural component of the myelin membrane. Hexadecanoic acid (palmitate) is a part of the fatty-acid substrate that supports myelin lipid biosynthesis (41). Choline contributes to phosphatidylcholine and sphingomyelin metabolism (42). Magnesium supports oligodendrocyte function and myelination markers in preterm models (43). In addition is has been shown how antenatal magnesium sulfate is associated with more mature white matter structure at term-age (higher FA), (44). Niacin supports NAD-dependent energy metabolism (45). Finally, Docosapentaenoic acid (DPA) is an intermediate in the biosynthetic pathway to DHA (46).

**Figure 1.**
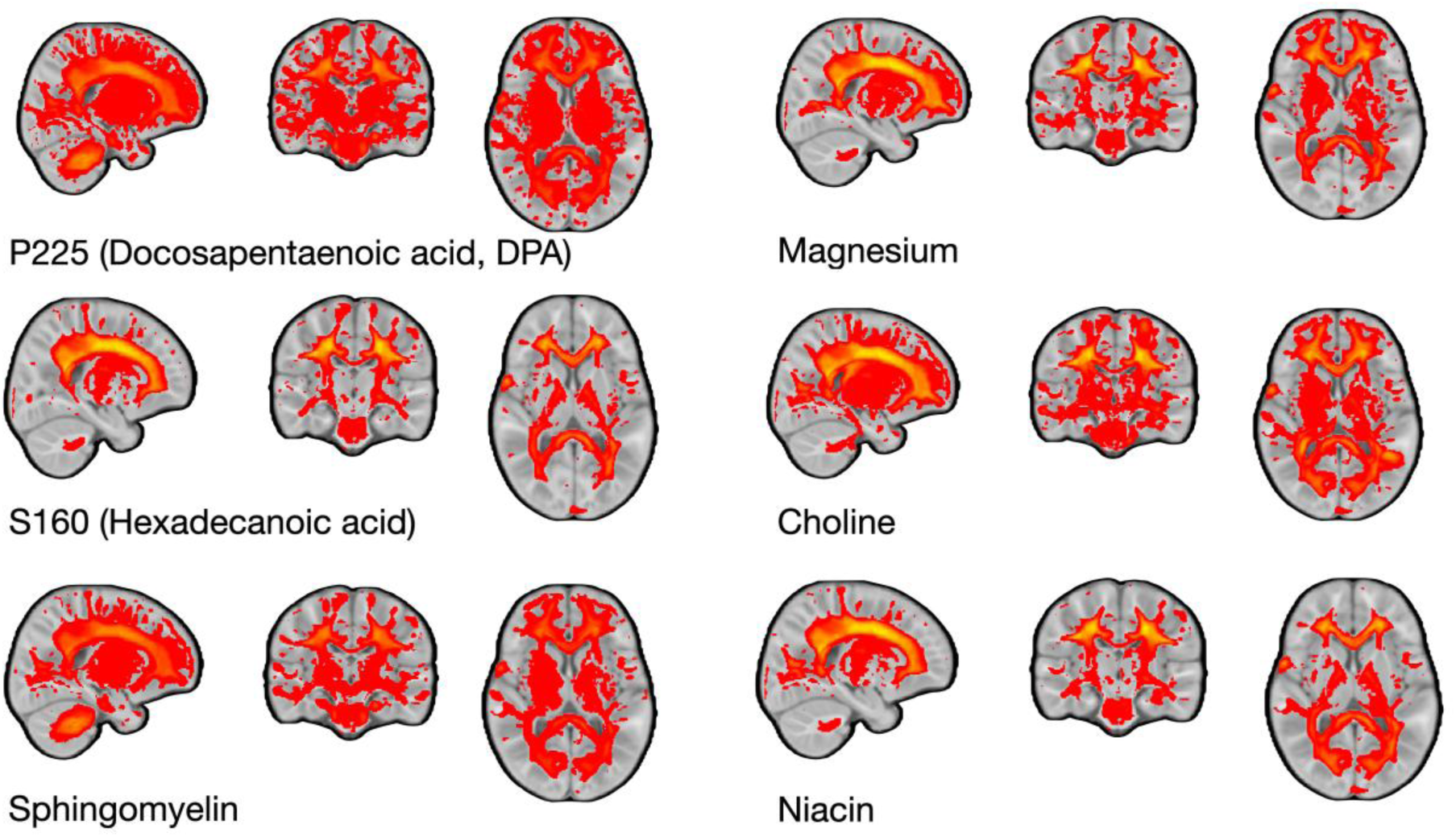
Nutrients for myelination. Brain regions with significant positive associations between white matter MWF and nutrient intake.

**Figure 2.**
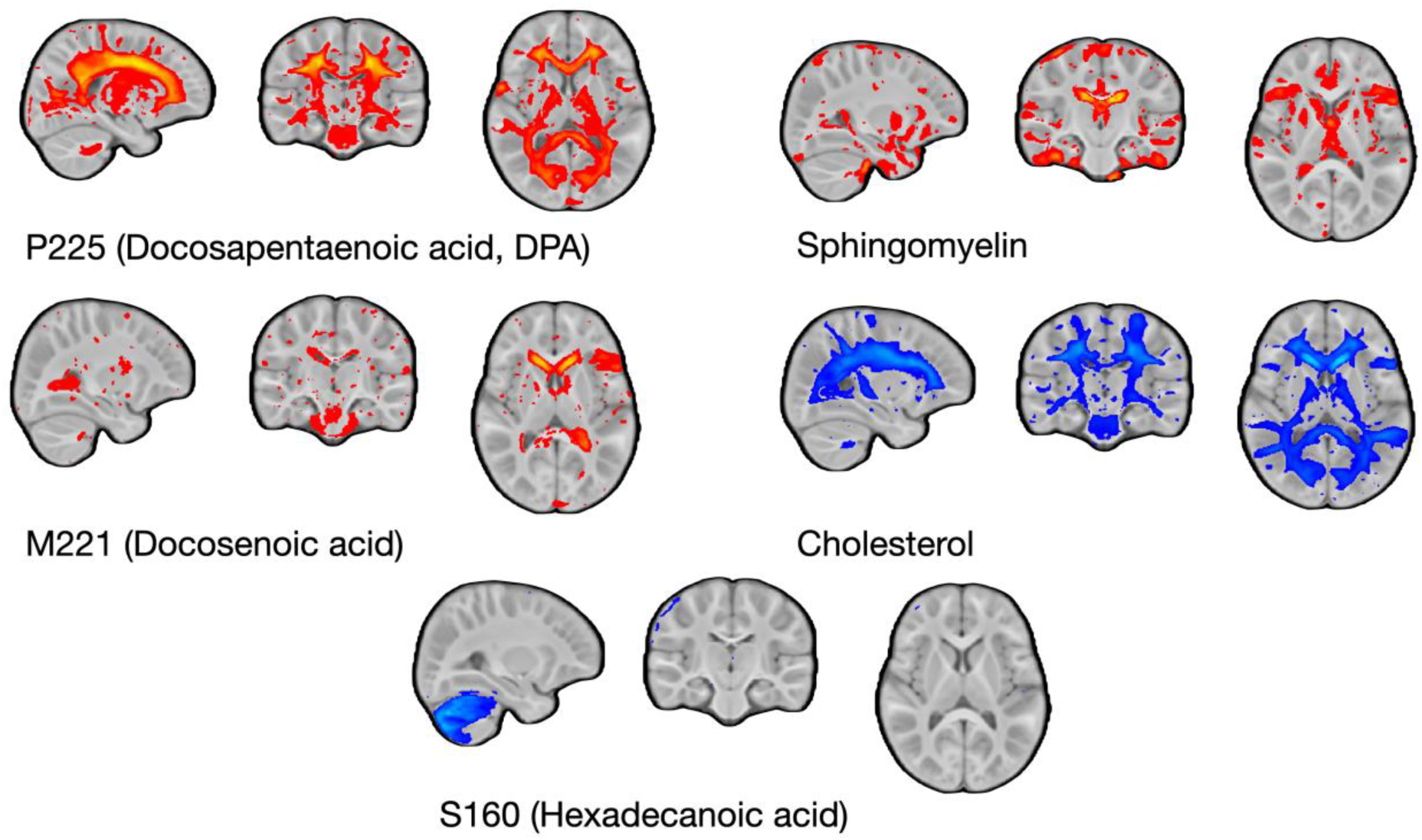
Nutrients for brain FA. Brain regions with significant associations between white matter FA and nutrient intake. Positive associations are shown in red, and negative associations are shown in blue.

Nutrients associated with brain FA included sphingomyelin, and docosenoic and docosapentaenoic acid (DPA) showing positive association, while cholesterol and hexadecanoic acid (palmitate) were found to have a negative association with FA. As mentioned previously, sphingomyelin is major myelin component that supports oligodendrocyte differentiation and myelination (47). Docosenoic acid and DPA are long-chain fatty acids with roles in membrane structure and DHA biosynthesis, respectively (48). On the other hand, cholesterol is an essential myelin component but its excess or dysregulation might impair axonal integrity and FA, via indirect vascular and inflammatory effects that promote oxidative stress, endothelial dysfunction and BBB permeability (49–51).

Similarly palmitate (hexadecanoic acid) as a saturated fatty acid can induce neuronal insulin resistance and mitochondrial dysfunction when elevated, potentially disrupting white matter microstructure (52).

### Associations Between Nutrition Intake and Cognitive Development

Nutrients with significant intake associations with cognitive (FISQ) and learning scores (AAB MC and AAB LWR) are summarized in **Table 2**. For cognitive skills (FISQ) significant associations were observed for folate, iron, sphingomyelin, vitamin B12, hexadecenoic acid (palmitoleic acid, commonly known as omega-7), eicosenoic acid, octadecenoic acid, DHA, eicosatetraenoic acid, hexanoic acid, and hexadecanoic acid (palmitic acid). For AAB math skills (MC) and reading (LWR) the same nutrients were significant, expect DHA and hexanoic acid. Overall the most consistent associations across all three outcomes were folate, iron, sphingomyelin, vitamin B12, palmitoleic acid, octadecenoic acid, eicosenoic acid, DHA, and palmitic acid. Most of these nutrients have been previously associated with learning and cognition, based on the roles they play in one-carbon metabolism, oxygen transport and neurodevelopment, myelination, synaptic function, and membrane lipid synthesis (41, 46–50). Specifically, folate and vitamin B12 support one-carbon metabolism and have been linked to cognitive performance, iron is also essential for brain development and learning, sphingomyelin as previously mentioned is paramount for myelination and cognitive maturation, and DHA supports neuronal membrane function and development (58,59).

**Table 2.** Nutrients for IQ and learning. Summary of nutrients that showed a significant association between intake and cognitive score, controlling for total calories, age, biological sex, and maternal education.

| Nutrient | p value FSIQ | p value (AAB MC) | p value (AAB LWR) |
| --- | --- | --- | --- |
| FOLATE | 0.022580 | 0.006769 | 0.042188 |
| IRON | 0.002733 | 0.011280 | 0.013125 |
| SPHINGOMYELIN | 0.038954 | 0.005584 | 0.01843 |
| VITAMIN B12 | 0.003414 | 0.024607 | 0.002685 |
| M161<br>(Hexadecenoic acid) | 0.008991 | 0.010512 | 0.039180 |
| M181<br>(Octadecenoic acid) | 0.019014 | 0.010015 | 0.031341 |
| M201<br>(Eicosenoic acid) | 0.033866 | 0.027761 | 0.042455 |
| P204<br>(Eicosatetraenoic acid) | 0.004869 | 0.004594 | 0.015584 |
| DHA<br>(Docosahexaenoic acid) | 0.026333 | 0.014237 |  |
| S060<br>(Hexanoic acid) | 0.049823 | 0.050372 |  |
| S160<br>(Hexadecanoic acid) | 0.02876 | 0.021380 | 0.045738 |

Overall, we find important overlap between nutrients associated with cognitive performance and some previously associated with brain white matter myelination, including sphingomyelin, hexadecenoic acid, and octadecenoic acid.

### Mediation Analysis linking Nutrition, White Matter Microstructure, and Cognitive Development

Putting results from the previous association analyses together into a formal mediation model, and restricting analysis to brain regions shown in Figure 1 and 2 and nutrients shown in Table 2, we find multiple brain regions in which MWF positively mediates the relationship between nutrition and FSIQ (**Figure. 3**) and AAB MC and LWR (**Figure. 4**). We also find regions where FA positively mediates the relationship between nutrition and FSIQ (**Fig. 5**). White matter FA was not found to significantly mediate nutrition-cognitive relationships for AAB MC or AAB LWR.

**Figure 3.**
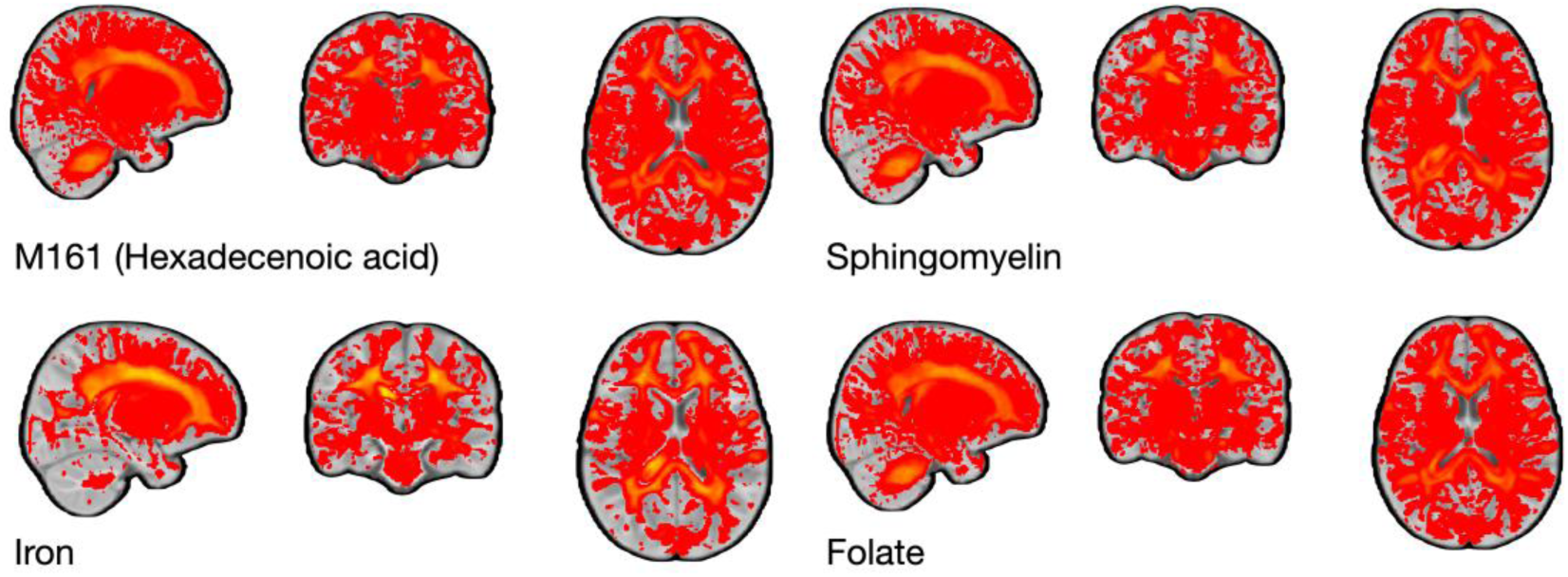
Myelination and IQ. Brain regions in which myelination (MWF) significantly (and positively) mediate the relationship between nutrient intake and WISC-V FSIQ cognitive measures.

**Figure 4.**
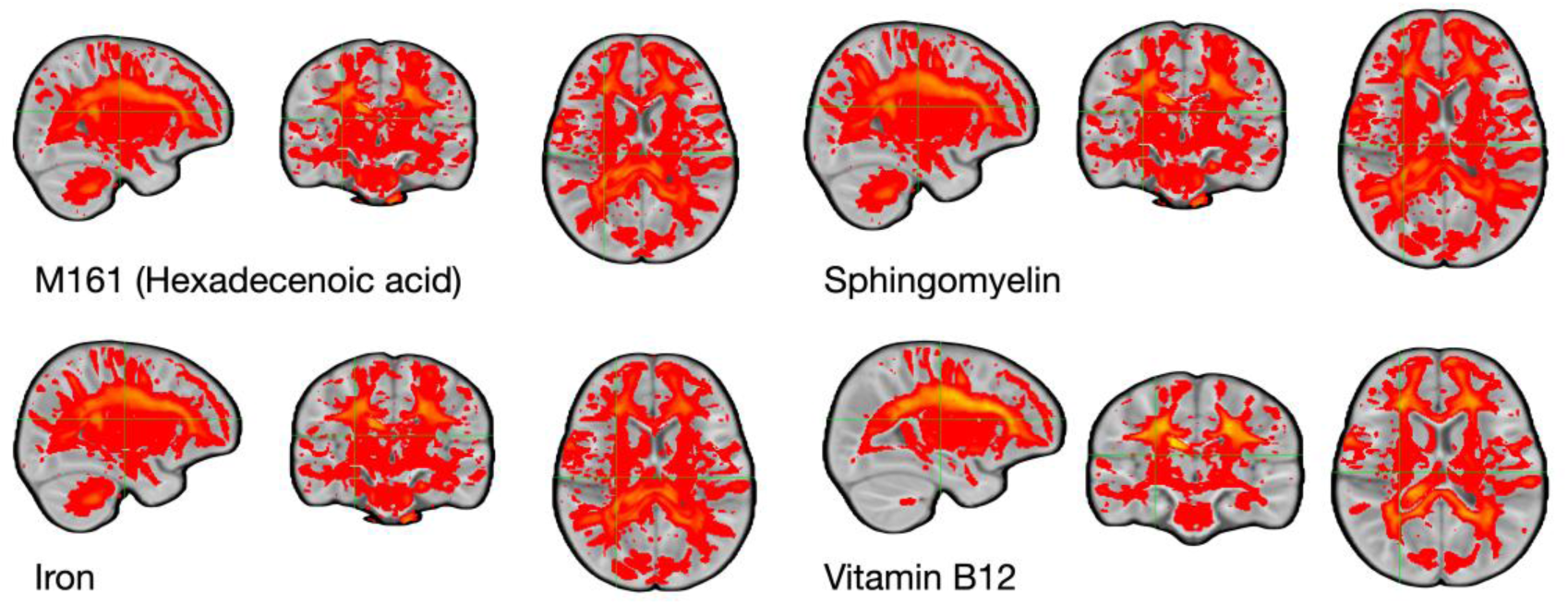
Myelination and learning. Brain regions in which myelination (MWF) significantly (and positively) mediate the relationship between nutrient intake and AAB MC and LWR learning measures.

**Figure 5.**
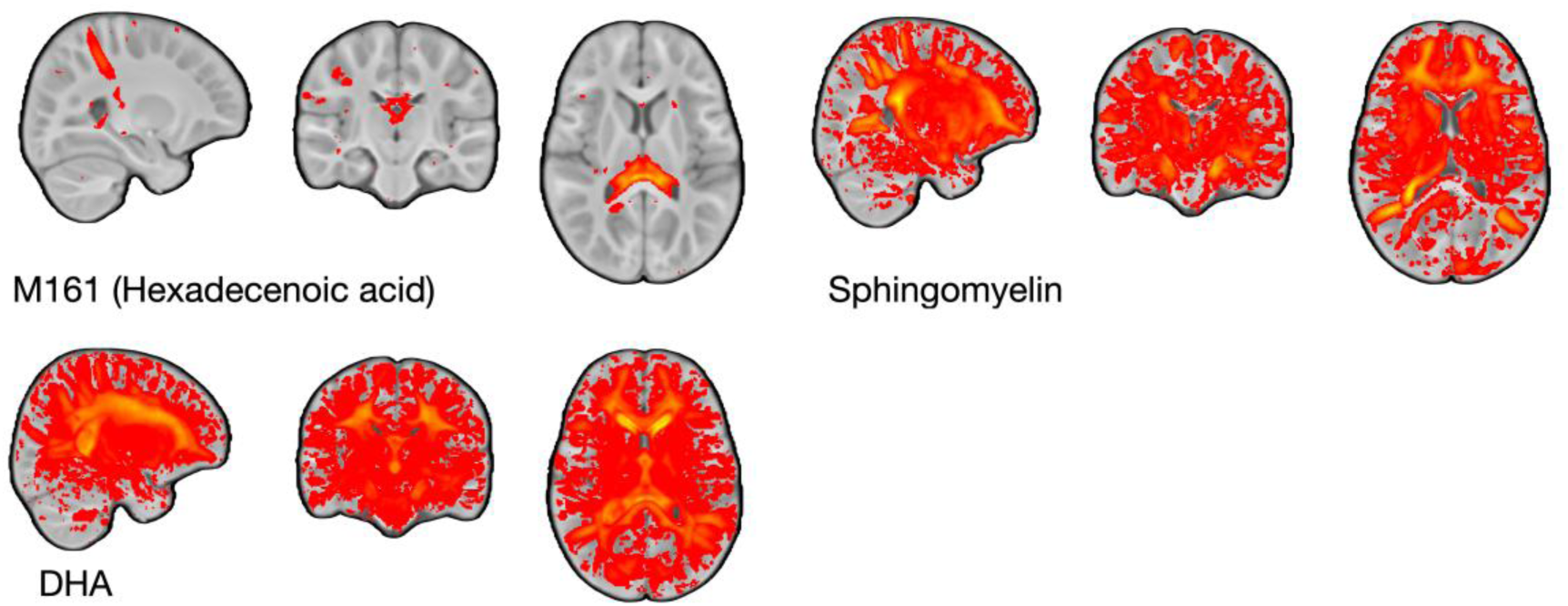
FA and IQ. Brain regions in which white matter microstructure (FA) significantly (and positively) mediate the relationship between nutrient intake and WISC-V FSIQ cognitive measures.

### Mediation Analysis linking Nutrition, Structural Connectivity, and Cognitive Development

Building on the microstructure analysis (MWF and FA), mediation analyses highlighted the importance of nutrients, including folate, vitamin B12, magnesium, iron, as well as lipids and fatty acids, including DHA, ARA, cholesterol, sphingomyelin, and hexadecenoic acid, across the measured cognitive outcomes (Figs. 6-8). These findings are consistent with the mediation outcomes described above and support the importance of structural development, specifically myelination, as a microstructural basis for structural connectivity.

**Figure 6.**
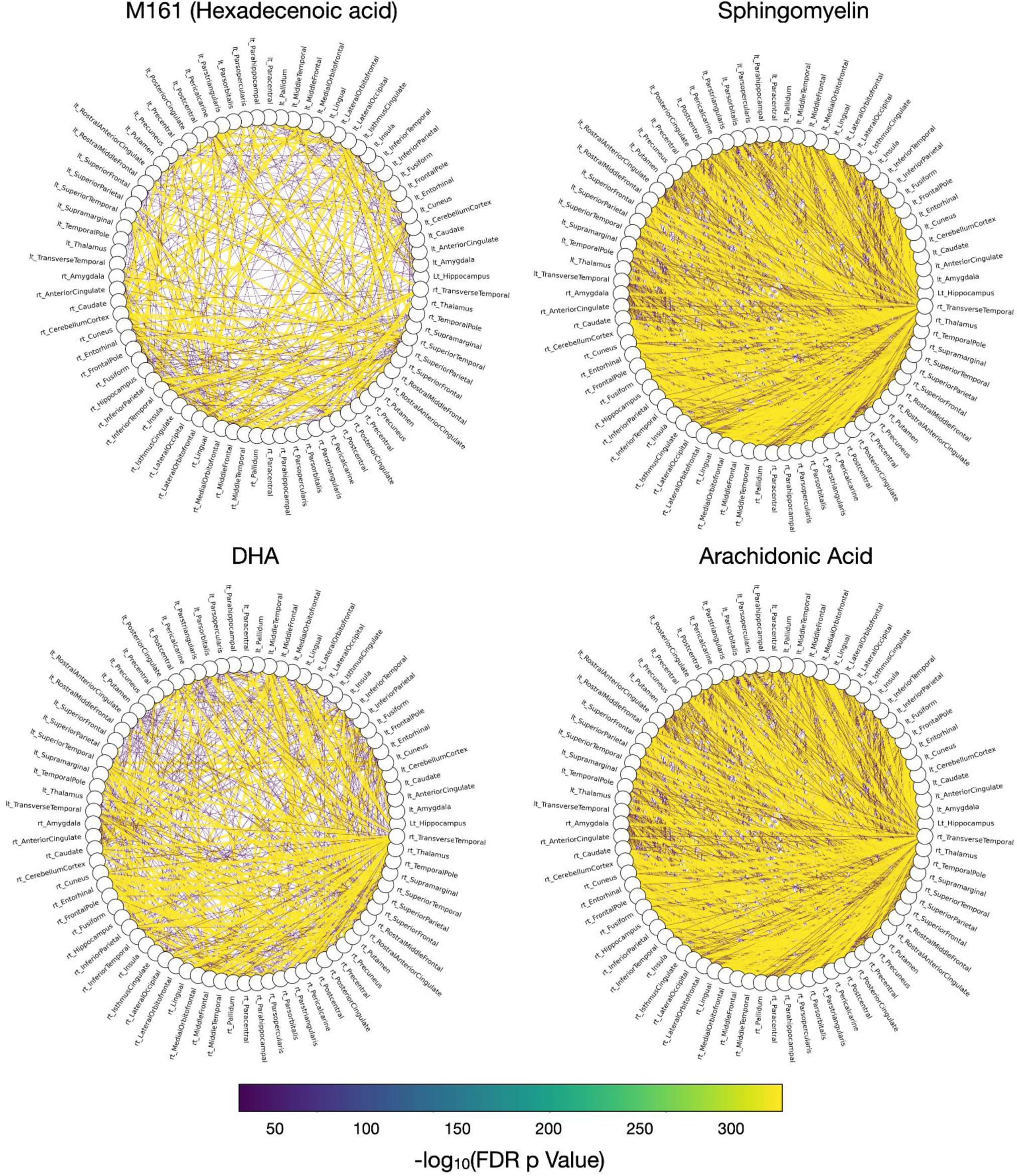
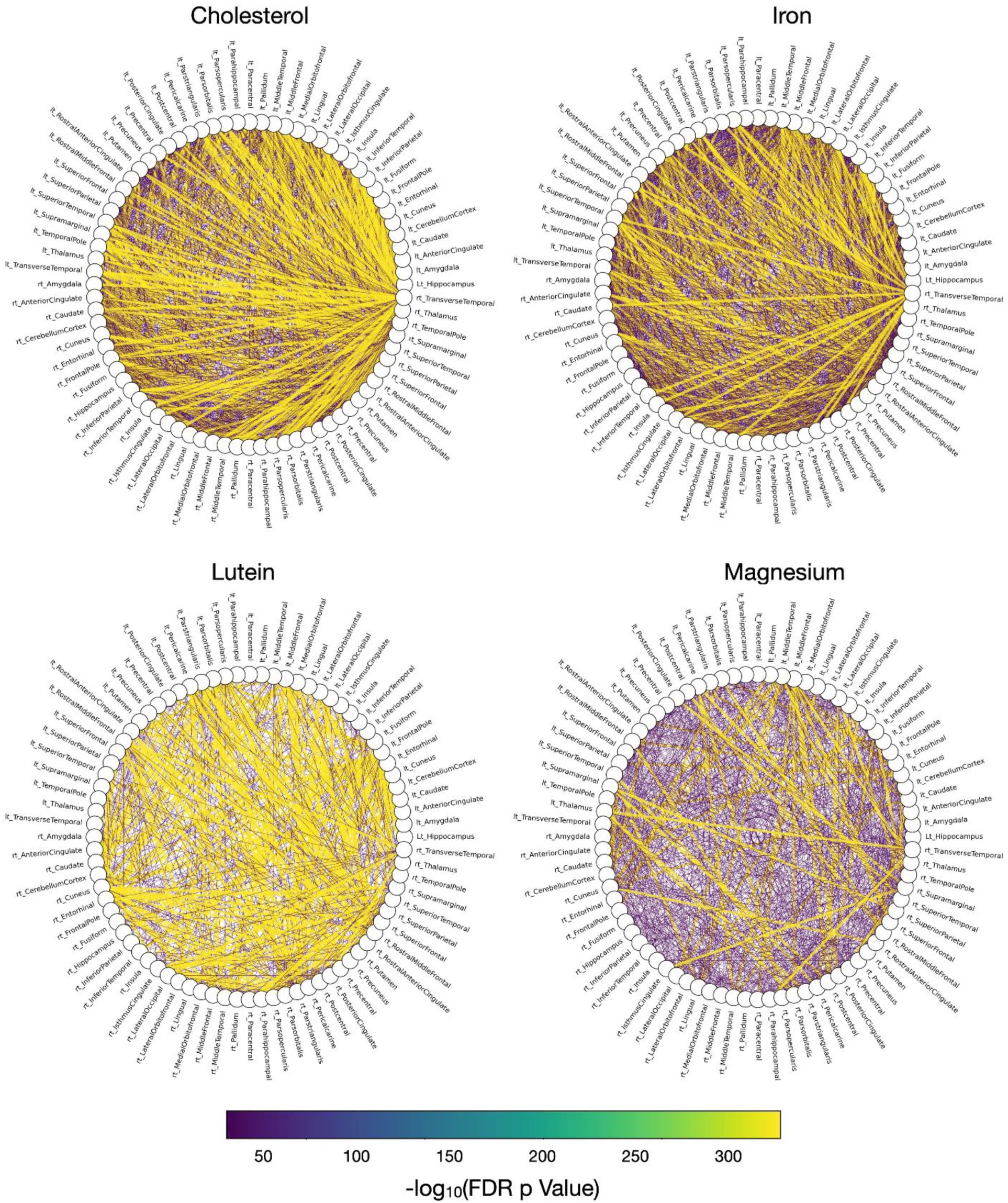
Structural connectivity and IQ. Connectivity diagrams highlighting structural connections that mediate the relationship between nutrient intake and FSIQ.

**Figure 7.**
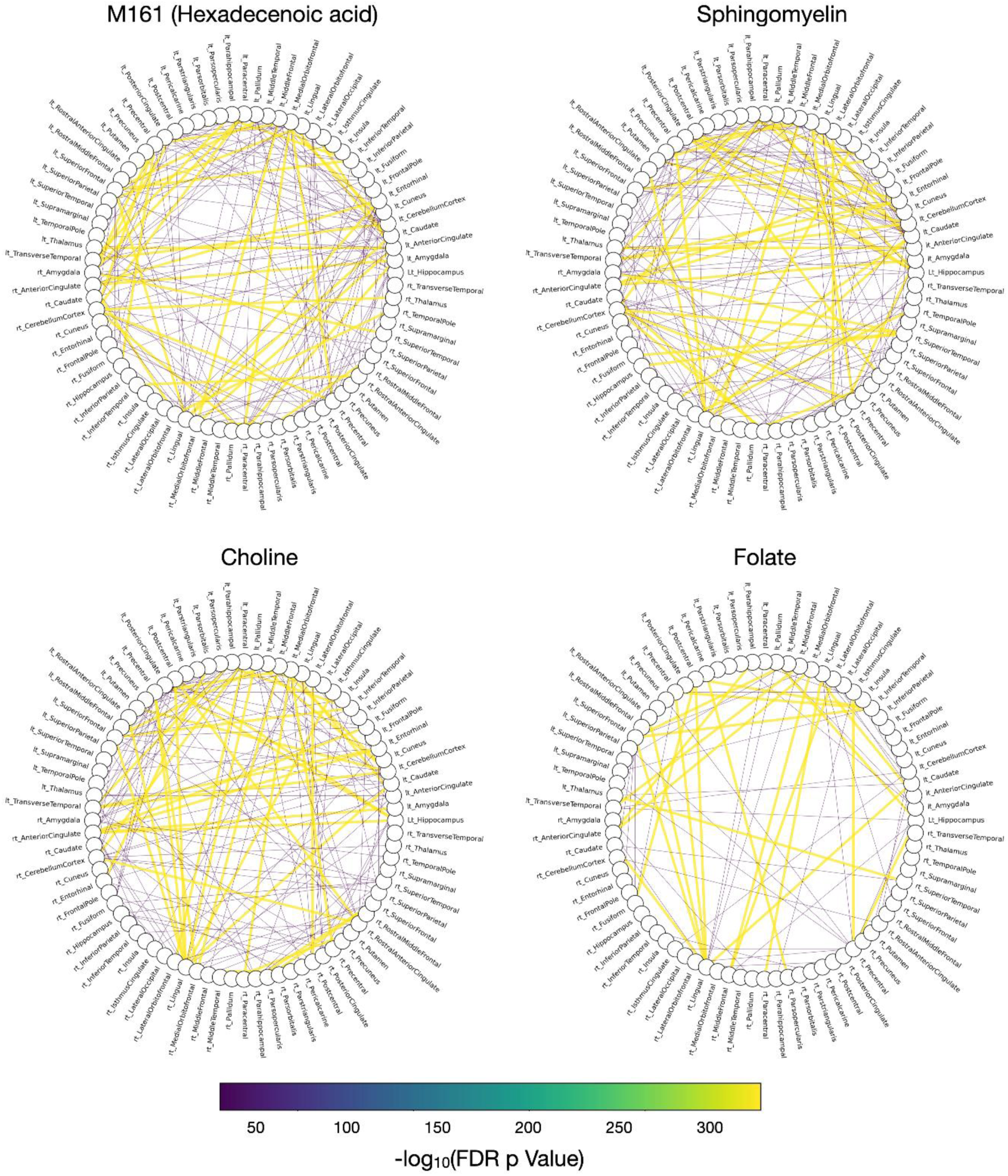
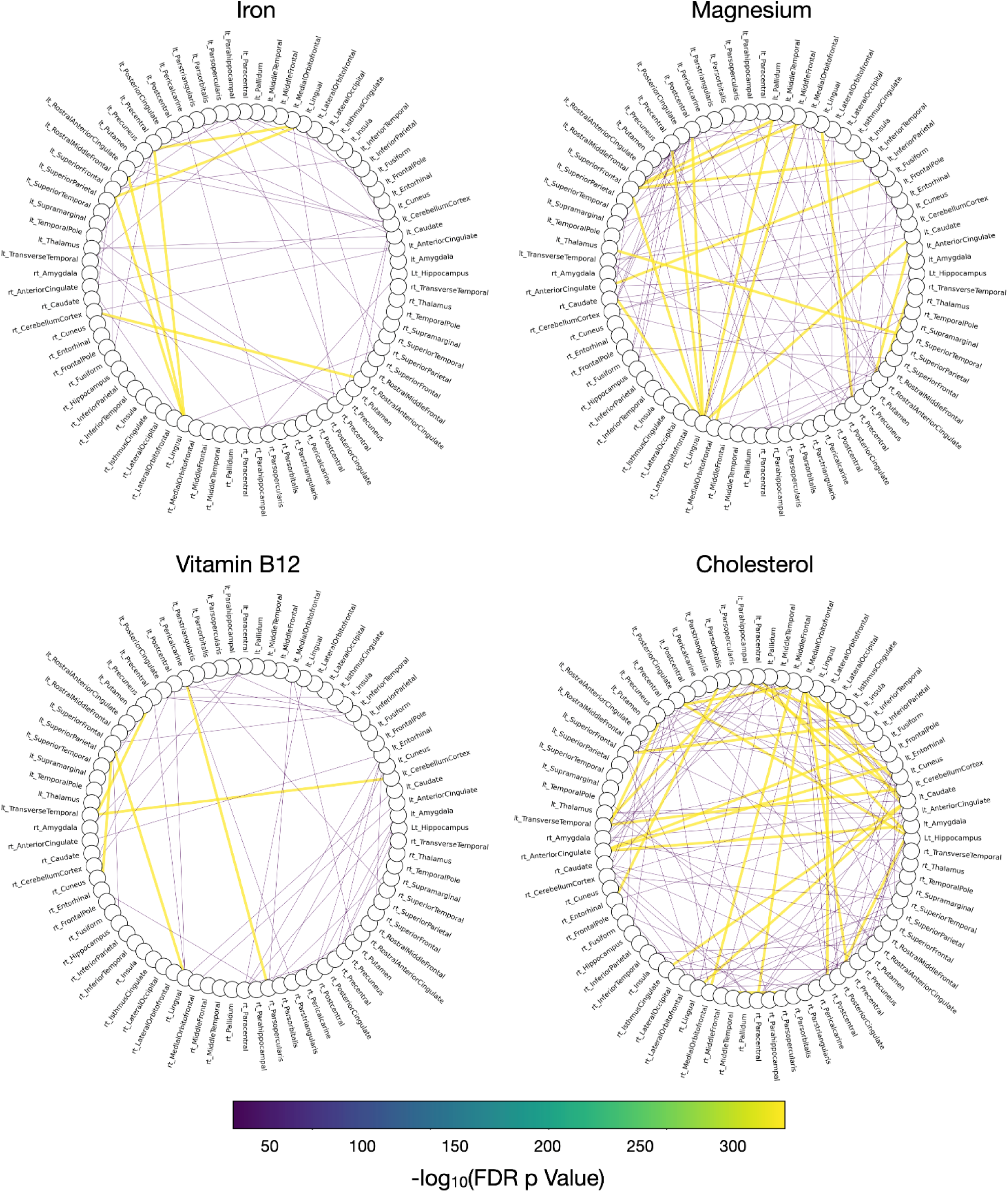
Structural connectivity and math comprehension. Connectivity diagrams highlighting structural connections that mediate the relationship between nutrient intake and Math Comprehension.

**Figure 8.**
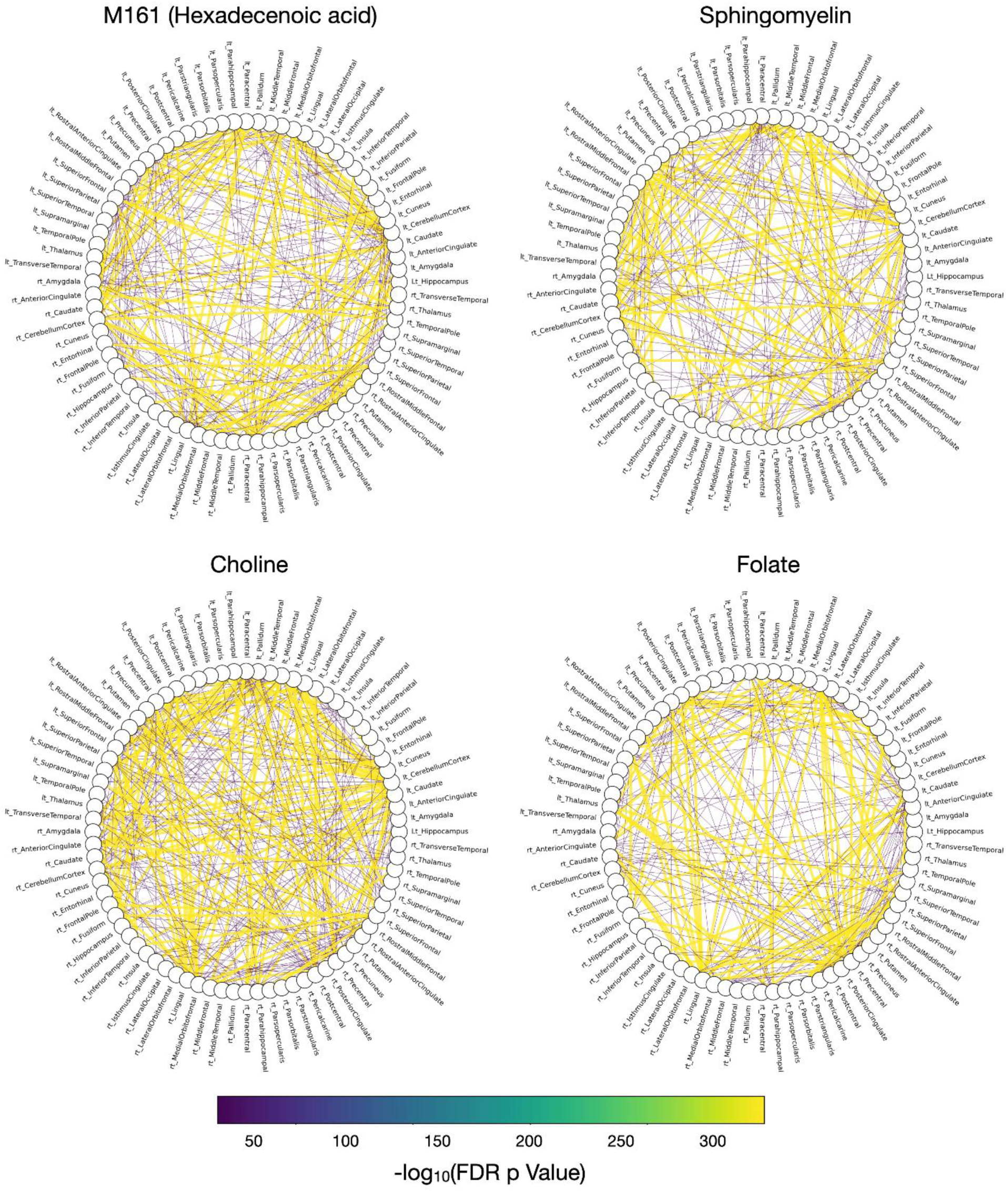
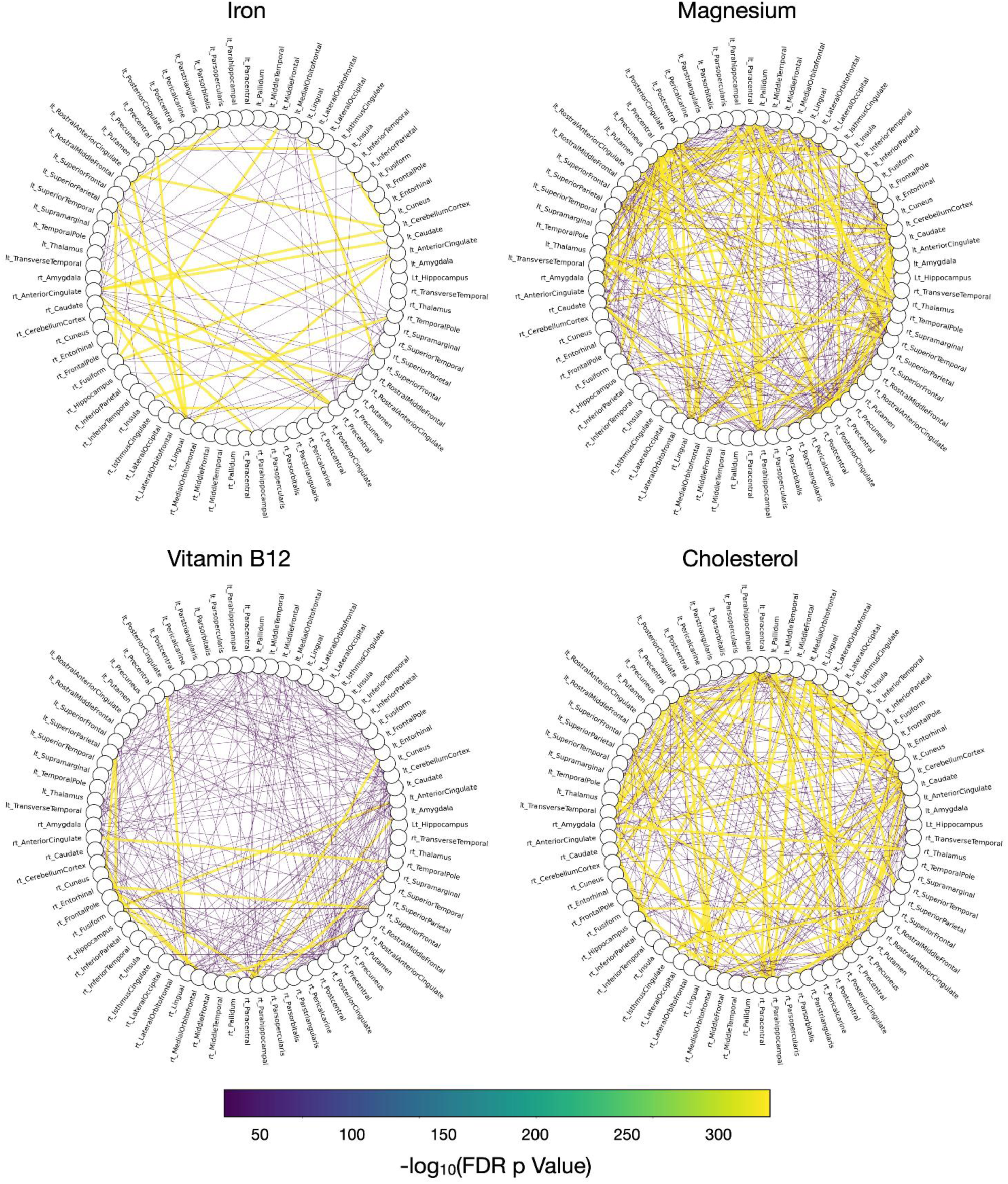
Structural connectivity and reading. Connectivity diagrams highlighting structural connections that mediate the relationship between nutrient intake and Letter-Word Recognition.

In addition, to determine if nutrient intake impacted the brain uniformly or if the effects were limited to specific brain ‘hubs’, we quantified degree centrality and degree count as the proportion (and absolute number) of significant nutrient-associated connections for each brain region. Regions with elevated degree centrality were interpreted as network hubs exhibiting widespread covariance relationships across the brain.

For general intelligence, we found that cholesterol, sphingomyelin, cholesterol, DHA, ARA, hexadecenoic acid (M161), and iron had broad impact across the brain, with all regions have a degree centrality greater than 0.2 (degree count > 15). Lutein and magnesium had more focal effects.

### Mediation Analysis linking Nutrition, Structural Connectivity, and Academic achievement

Similarly academic achievement outcomes (both math comprehension and word-letter recognition), the effect of nutrition on connectivity was more focused on specific regional hubs. For both academic outcomes, this included left hemisphere precentral and paracentral gyrus, anterior cingulate, amygdala, hippocampus, and superior frontal gyrus; and bilateral lingual gyrus and cerebellar cortex. Figure 8 illustrates the significant hubs for AAB math comprehension, and Figure 9 the significant hubs for AAB reading.

## DISCUSSION

Multiple studies have highlighted the importance of nutrition and specific nutrients for early brain and cognitive development. However, far fewer studies have extended this work into later childhood and beyond specific deficiency states (e.g., iron-deficiency) and in typically developing children. Recent systematic review showed how only a small number of studies in school-age have incorporated brain imaging (18). To the best of our knowledge, this is the first study to combine detailed nutritional and cognitive assessment with extensive structural imaging to examine associations between nutrient intake, white matter microstructure and connectivity, learning, and cognition in a large cohort of neurotypical school-age children.

We first examined direct associations between diet-derived nutrients and three domains: white matter microstructure (MWF and FA), cognitive performance, and academic achievement. We then applied mediation analyses to assess whether white matter microstructure and connectivity helped explain associations between nutrient intake and cognitive and learning outcomes. Together, these analyses suggest that nutritional intake during childhood may relate not only to the organization of white matter development, pathways and connectivity, but also to cognitive and academic performance.

Several nutrients showed robust associations with white matter microstructure (MWF and FA) and cognition, including choline, sphingomyelin, docosapentaenoic acid, hexadecanoic acid, magnesium, niacin, and other fatty acids. These associations are biologically plausible given the previously established roles of these nutrients in membrane synthesis, lipid metabolism, and myelination (39,60,61). In particular, choline and sphingomyelin are directly implicated in myelin formation and maintenance, while fatty acids such as docosapentaenoic acid and hexadecanoic acid may support the lipid supply required for white matter growth (42,55,62,63). The pattern of findings supports the idea that nutritional substrates involved in membrane assembly remain relevant across childhood (64).

Our findings extend this literature by showing that nutrient-white matter associations are detectable beyond infancy, into school age. Docosapentaenoic acid and DHA were also associated with white matter integrity, consistent with the importance of long-chain omega-3 fatty acids for brain lipid composition and neurodevelopment (65).

The effects of hexadecanoic acid and cholesterol were more complex. Both are essential components of lipid metabolism and myelination, yet their associations with white matter structure differed across imaging measures. This likely reflects the fact that myelin water fraction and fractional anisotropy capture distinct features of white matter organization and may respond differently to lipid availability or metabolic imbalance (66). Rather than indicating a simple beneficial effect, these findings suggest that the relationship between lipid intake and white matter is nuanced and may depend on the specific nutrient, brain region, and microstructural metric (67).

Mediation analyses further showed that several nutrients were linked to cognitive and academic outcomes through white matter pathways. In particular, folate, iron, sphingomyelin, DHA, palmitoleic acid (hexadecenoic acid), and choline emerged as consistent contributors across models. This pattern supports a model in which childhood nutrition influences cognition partly through its effects on white matter development (68). More broadly, these results suggest that nutritional factors relevant to myelination, membrane metabolism, and neural connectivity may continue to shape brain organization and cognitive function throughout childhood.

Future studies should investigate these nutrient patterns at a more mechanistic level to better understand how combinations of nutrients influence brain development, metabolism, learning, and cognitive performance. In particular, in vitro and in vivo models could help clarify whether the observed associations reflect direct effects on myelination, lipid metabolism, oligodendrocyte function, synaptic development, or broader neurodevelopmental pathways. Such work would also help determine whether the effects of individual nutrients differ from those of nutrient mixtures or dietary patterns, and whether specific combinations have additive, synergistic, or even opposing effects on brain maturation and cognitive outcomes.

## CONCLUSIONS

The present study examines the relationship between daily nutrition, white matter development and connectivity, and cognitive and learning outcomes in a large cohort of school-age. Using a data-driven association and mediation analyses, we identified key nutritional components linked to white matter microstructure and connectivity, cognition, and learning with lipids emerging as especially relevant nutritional components. Specifically, sphingomyelin, DHA, and palmitoleic acid (omega-7) were recurrent components mediating the effects between nutrition and cognitive and learning outcomes. These findings support the idea that brain development in childhood is shaped and supported by multiple nutrients interacting rather than by single components. Future studies should test the synergistic and cumulative effects of these nutrients in relevant dietary interventions.

## LIMITATIONS

Several limitations need to be considered. First, the current study is observational in nature and hence cannot establish causality, even though mediation modelling clarifies possible plausible pathways. Second, dietary intake was based on the mean across 3-days of recall-based assessment, which might be affected by reporting error. Finally, the sample was restricted to typically developing children, so current findings cannot generalise to clinical or at-risk populations and to other developmental stages (i.e., infancy).

## Data Availability

All data produced in the present study are available upon reasonable request to the authors.

https://echochildren.org/dash/

## ACKNOWLEDGMENTS & CONTRIBUTIONS

The authors thank all study participants for their interest and willingness to contribute to the study. Furthermore, the authors thank the clinical site teams and the analytical teams for their invaluable contributions. DB, JH, IS, and FM are employed by Société des Produits Nestlé S.A. DB and SD conceptualised the study design and wrote the manuscript. FM, CER, and SD performed the analyses. JH and IS provided their feedback on nutritional interpretation. All the co-authors participated in manuscript drafting and revision.

